# Determining the Origins of Repeat *Trichomonas vaginalis* Infections Using Clinical Versus Genotype-Informed Criteria

**DOI:** 10.1101/2022.01.31.22268862

**Authors:** Heather Larkin, Martina Bradic, Norine Schmidt, David H. Martin, Jane M. Carlton, Patricia J. Kissinger

**Author notes:** **Corresponding author:** Patricia J. Kissinger, PhD, 1440 Canal St. #8318, New Orleans, LA 70112.

## Abstract

**Background:** High rates of repeat infections post-treatment are reported in women infected with *Trichomonas vaginalis* (TV). Determining the origin of repeat infections is generally limited to clinical queries of adherence to treatment and sexual exposure. The purpose of this study was to add micro-satellite (MS) genotype data to classification criteria for origin of repeat TV infection, and examine if the addition of TV genotype changes classification as treatment failure, re-infection, or new infection

**Methods:** Women were enrolled at clinics in Birmingham, AL; Jackson, MS; and New Orleans, LA as part of a randomized clinical trial comparing single-dose (2 g) and multi-dose (500 mg twice daily x 7 days) metronidazole (MTZ) treatment regimens. Participants provided vaginal swabs and completed a behavioral audio-computer assisted self-interview (ACASI). TV specimens were genotyped at 11 microsatellite (MS) loci. Women with repeat TV infections at TOC, were classified as treatment failure, re-infection or new infections using behavioral and genotype data; classifications were compared.

**Results:** Data were available for 45 women. Genotype concordance was defined as <4 MS loci different and genotype discordance was defined as ≥ 4 MS loci different. Clinical criteria vs. genotype-informed criteria classifications were treatment failure (66.7% vs 64.4%) re-infection (26.7% vs. 17.8%) and new infections (6.7% vs. 17.8%) respectively; Bowker’s test of symmetry had Χ^2^=16.00 p=0.0011, indicating differences in results.

**Conclusions:** The majority of women, using either criteria, were classified as treatment failure. Clinical assessment may overestimate reinfections and underestimate new infections. Patient counseling should be adapted accordingly.

**Summary:** To more precisely determine the origin of repeat *Trichomonas vaginalis* infection, we compared genotype microsatellite size polymorphism data to clinical criteria and found that clinical data overestimated reinfection and underestimated new infections. Patient counselling should consider new partners.

## Background

*Trichomonas vaginalis* (TV) is the most common curable sexually transmitted infection (STI). There is an estimate prevalence of 5.3% TV in women globally, and 3.7 million cases in the United States[1]. Infection with this haploid, flagellated protist is associated with adverse outcomes for pregnant women and their offspring, increased risk of HIV transmission, inflammation of the urogenital tract, and cancers of the cervix and prostate [2, 3]. The burden of this disease is not distributed equally in the population. *T. vaginalis* was twelve times higher in non-hispanic black females (9.6%; 95% CI, 7.3%–12.5%) compared to non-Hispanic white females (0.8%; 95% CI, 0. 4%–1.5%) in NHANES nationally representative sample from 2013-2016 [4, 5].

Repeat infection after treatment occurs in 5-33% of women [6-8]. Drug resistance, poor adherence to treatment, or treatment regimen recommended can cause repeat infection via failure of metronidazole (MTZ) treatment to eliminate all parasites in the host [9, 10]. Reinfection by baseline partners who are infected with the original parasite, a new parasite strain from a baseline partner, or infection from a new partner may also cause repeat infection [11].

Methods to determine the origin of repeat infection are often limited to a clinical questionnaire which records treatment adherence, sexual exposure to partners during and after MTZ treatment, and barrier use. Our group has previously reported more than 50% of repeat infections are due to treatment failure in both HIV-infected and uninfected women, and repeat infection from unprotected sexual exposure accounts for a minority of cases [8].

Short tandem repeats (STRs), or microsatellites (MS) in the genomes of pathogens are sequences of 3-6 base pairs that repeat a variable number of times between different strains strains of the same species[12]. Panels of MS markers have been developed for use in TV[13], including a panel of 21 MS markers used in studies of population genetics and TV in the context of the bacterial microbiome. The unique profile of repeats at MS loci can provide a genotype with which to differentiate the strain present at baseline and test of cure (TOC). In our previous study in HIV-positive women, repeat infections were on account of 36% treatment failure, 6% incomplete treatment, 21% re-infection by baseline partners, 9% by exposure to a new partner, and 27% possible new infections [14] using 11 microsatellite loci from this panel at baseline and TOC [13].

The purpose of this study was to evaluate the origins of repeat TV infection with standard clinical criteria and with clinical criteria informed by the TV MS genotype at baseline and four weeks post-treatment TOC in HIV-uninfected women from the southern United States. We defined the algorithm to add MS genotype data to classification criteria for origin of repeat TV infection, determined the threshold of MS repeats used to define TV strain as concordant or discordant, and examined if the addition of TV genotype changes the proportion of cases classified as treatment failure, re-infection, or new infection.

## Methods

### Study design

This study was part of a phase III clinical randomized controlled trial (RCT). Methods have been described elsewhere [9]. Briefly, subjects were randomized 1:1 to receive the standard of care, a single dose of 2 g metronidazole (MTZ), or 500 mg MTZ twice a day for seven days. Those eligible to participate were female sex, English-speaking, age of majority, tested positive for TV, and willing to refrain from use of alcohol during and 24 hours after treatment. Women with HIV, pregnant, breastfeeding, incarcerated, medical contraindications to MTZ treatment, or treated with medications used to treat TV or bacterial vaginosis (BV) within the last 14 days were excluded. All participants were enrolled at sexually transmitted disease (STD) clinics in Jackson, MS, Birmingham AL, and New Orleans, LA. Study methods for the RCT are described elsewhere [9]. In brief, TV+ women enrolled in the study provided a vaginal swab for culture and metronidazole resistance testing. Participants answered a behavioral survey using audio-computer assisted self-interview (ACASI) at their enrollment and TOC visits. The TOC visit was scheduled four weeks post-treatment completion. The TOC survey included questions on medication adherence. At each visit, InPouch™ culture specimens were processed for genotyping after being identified as positive for TV. This study was conducted in accordance with approval of the Institutional Review Boards of Tulane University and all partner institutions.

### Culture Processing

The swab for TV diagnosis was placed in InPouch™ culture medium and incubated at 37°C. Laboratory technicians at the clinical sites microscopically evaluated presence of the trichomonads in the cultures per InPouch™ guidelines. Once the final read for TV was made, samples were processed for genotyping. The sample was maintained until a minimum of 50 parasites per field at 10x were visible. Samples were spun down, re-suspended in Diamond’s media with 10% DMSO and 10% horse serum. This suspension was aliquoted into two cryotubes, frozen at a minimum of -70 °C and batch shipped on dry ice to the Center for Genomics and Systems Biology in the Department of Biology at New York University (NYU).*DNA Extraction, Genotyping and Analysis*

At NYU, the parasites were cultured in Diamond’s media, and DNA from each sample extracted and genotyped using 11 MS loci as previously described[15]. Briefly, DNA extraction was performed using the UNSET buffer and phenol-chloroform extraction method and DNA resuspended in TE buffer, pH 8 (10mM Tris–HCl, pH 8, 0.1mM EDTA, pH 8.0) [13]. Genotyping PCR reactions were performed in duplicate, and discrepancies were verified with a third reaction. Size polymorphisms were measured by capillary gel electrophoresis on an ABI 3,130xl sequencer, and GeneMapper 4.0 (ABI, Foster City, CA) used to score MS allele sizes. All calls were manually edited to discard data from poorly amplified reactions and to ensure that proper allele calls were assigned [15].

### Clinical Classification

In the standard clinical classification algorithm, sexual exposure was defined as subject report of vaginal sex without a condom with at least one untreated baseline partner or at least one new partner between their enrollment and TOC visit. In the clinical classification algorithm, all those with no reported sexual exposure, or sexual contact with 100% sexual barrier use, were classified as treatment failure. Those with exposure to baseline partners with or without exposure to new partners were considered re-infection, and those with exposure to new partners only were classified as new infection.

### Analysis

The subset of women enrolled in the RCT that were included in this analysis were TV+ at both enrollment and TOC, had complete behavioral and genotype data and reported 100% adherence to the treatment regimen. Statistical analyses were performed using SAS 9.4 software (Cary, NC). The outcome for our third aim was classification of infection as treatment failure, re-infection, or new infection. We compared clinical criteria with genotypic criteria. We assessed the percent agreement not due to chance with Cohen’s kappa for reliability. We used Bowker’s test of symmetry to test whether the proportion of cases classified in the three categories changed with the two classification criteria.

## Results

From October 2014 to April 2017, 1028 female patients were assessed for eligibility for the RCT, of whom 623 were enrolled. Results of the RCT for MTZ treatment strategy have been reported elsewhere [9]. Among the 540 (87%) participants who returned for a follow-up visit, 80 experienced repeat TV infection. Seventy-eight of these participants provided clinical interview on sexual exposure, and 49 had samples that were genotyped. Samples contaminated by Candida or which failed in culture were not able to be genotyped. Four participants had missing or reported incomplete adherence. A total of 45 subjects were included in this analysis (Supplementary Table 1). The mean age was 31.5 years (range 19-58). All participants were black/African American. At baseline, the 45 participants reported a total of 79 partners, and 44.4% reported two or more partners. Thirty-three (73.3%) participants reported previous TV infection, and 43 (95.6%) reported at least one symptom. Most of the participants (75.6%) in this analysis were enrolled in Birmingham, AL. At baseline, no subjects had a mixed TV infection (defined as the detection of multiple alleles present at two or more MS loci in this haploid organism).

Two thirds (n=30) of women included in this analysis had been randomized to single-day 2 g MTZ treatment group, while 15 were in the seven-day MTZ group (Table 1). The percentage of outcomes that differed by classification criteria was similar by treatment arm (Supplementary Table 2). Two out of 45 samples showed low to moderate MTZ resistance. With clinical criteria, treatment failure accounted for 56.7% of recurrent infection in the single-day 2 g MTZ arm, and 33.3% of repeat infections in the seven-day treatment arm.

**Table 1.**
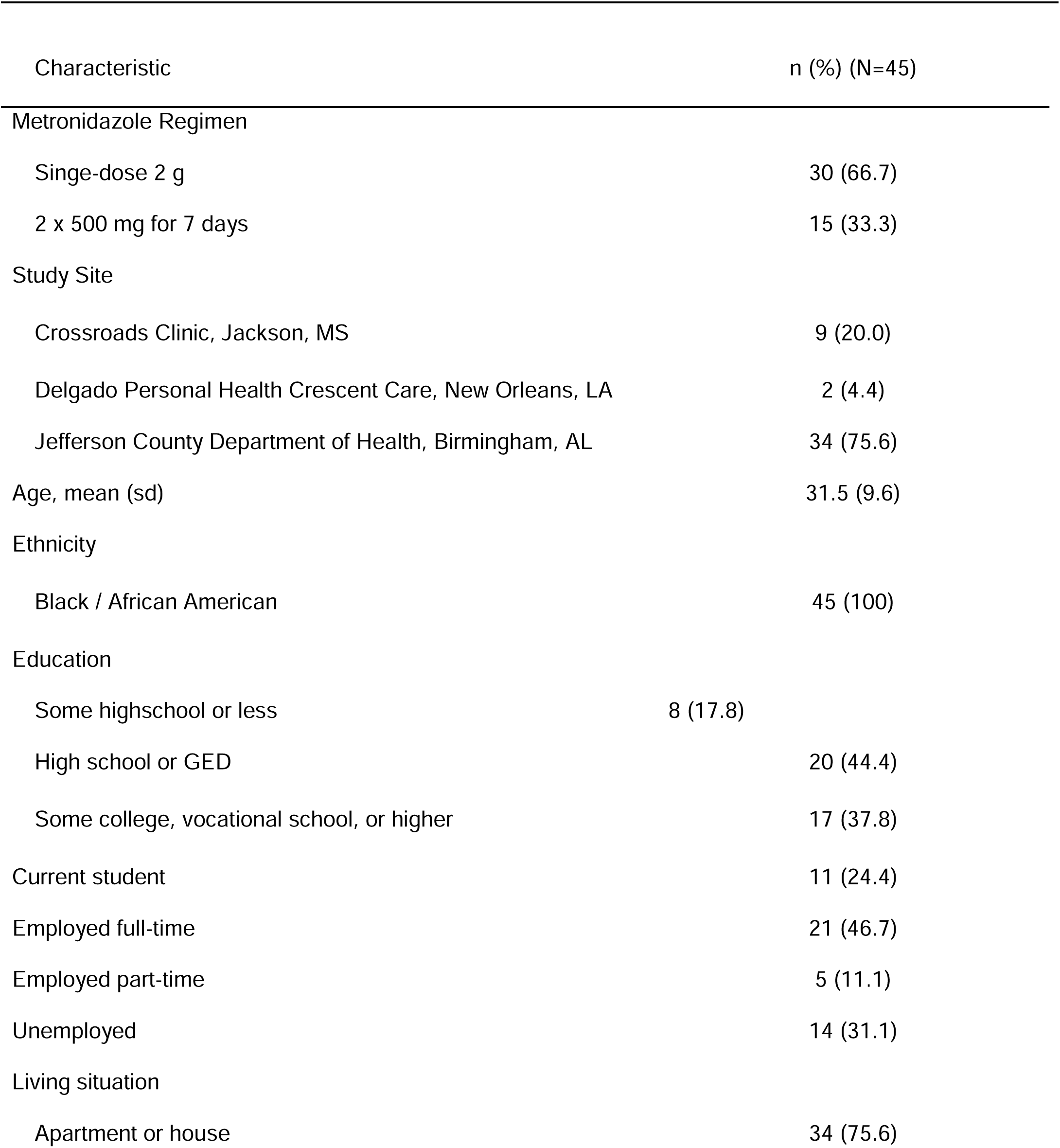
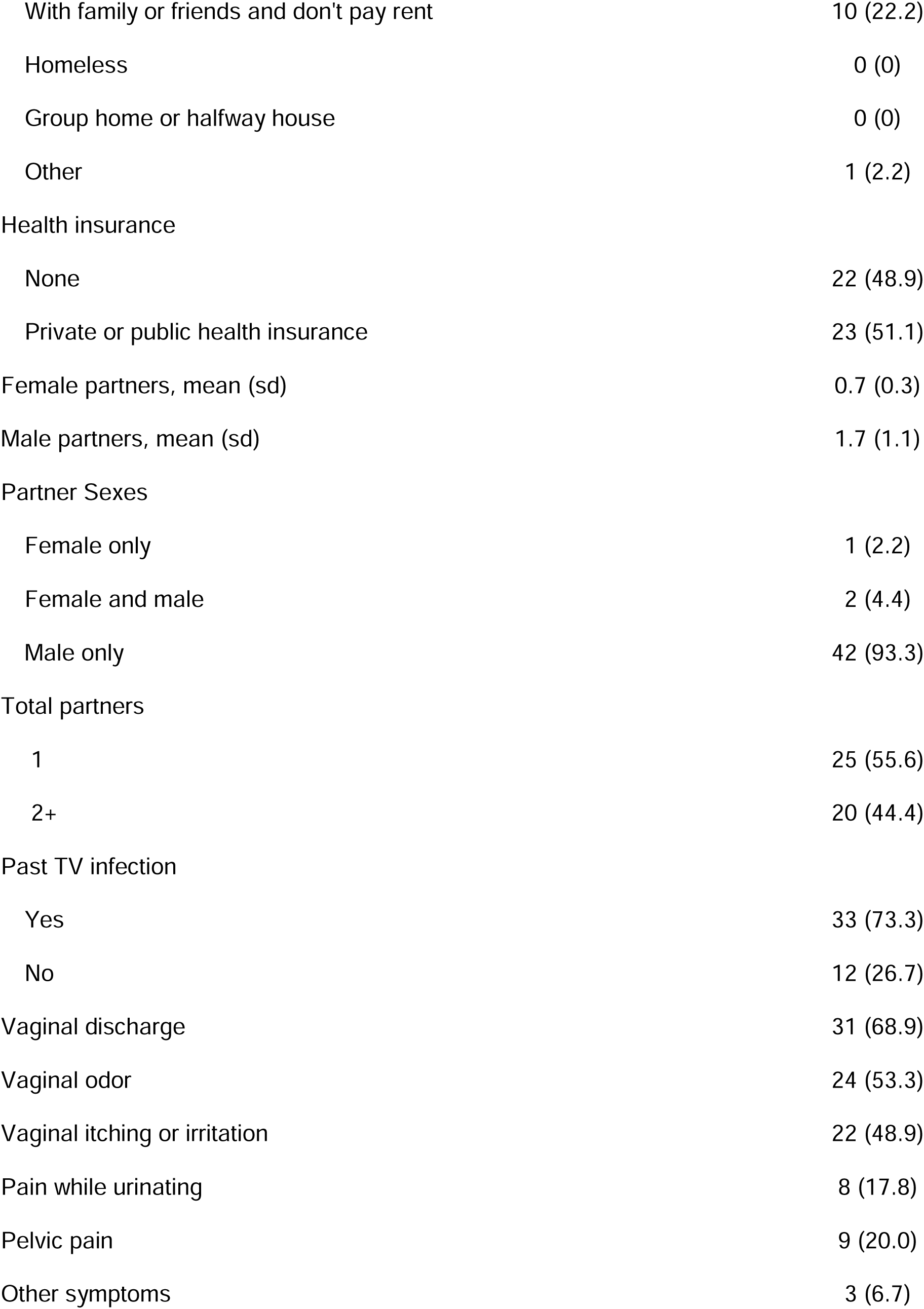
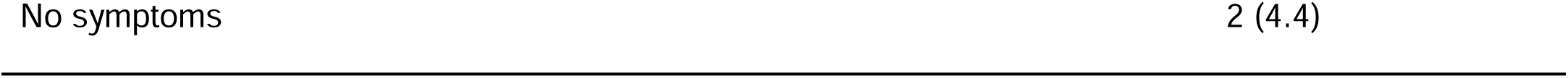
Background characteristics of participants

| Characteristic | n (%) (N=45) |
| --- | --- |
| Metronidazole Regimen |  |
| Single-dose 2 g | 30 (66.7) |
| 2 x 500 mg for 7 days | 15 (33.3) |
| Study Site |  |
| Crossroads Clinic, Jackson, MS | 9 (20.0) |
| Delgado Personal Health Crescent Care, New Orleans, LA | 2 (4.4) |
| Jefferson County Department of Health, Birmingham, AL | 34 (75.6) |
| Age, mean (sd) | 31.5 (9.6) |
| Ethnicity |  |
| Black / African American | 45 (100) |
| Education |  |
| Some highschool or less | 8 (17.8) |
| High school or GED | 20 (44.4) |
| Some college, vocational school, or higher | 17 (37.8) |
| Current student | 11 (24.4) |
| Employed full-time | 21 (46.7) |
| Employed part-time | 5 (11.1) |
| Unemployed | 14 (31.1) |
| Living situation |  |
| Apartment or house | 34 (75.6) |
| With family or friends and don't pay rent | 10 (22.2) |
| Homeless | 0 (0) |
| Group home or halfway house | 0 (0) |
| Other | 1 (2.2) |
| Health insurance |  |
| None | 22 (48.9) |
| Private or public health insurance | 23 (51.1) |
| Female partners, mean (sd) | 0.7 (0.3) |
| Male partners, mean (sd) | 1.7 (1.1) |
| Partner Sexes |  |
| Female only | 1 (2.2) |
| Female and male | 2 (4.4) |
| Male only | 42 (93.3) |
| Total partners |  |
| 1 | 25 (55.6) |
| 2+ | 20 (44.4) |
| Past TV infection |  |
| Yes | 33 (73.3) |
| No | 12 (26.7) |
| Vaginal discharge | 31 (68.9) |
| Vaginal odor | 24 (53.3) |
| Vaginal itching or irritation | 22 (48.9) |
| Pain while urinating | 8 (17.8) |
| Pelvic pain | 9 (20.0) |
| Other symptoms | 3 (6.7) |

At TOC, 30 of the 45 participants reported no sexual exposure (Figure 1), four reported exposure to baseline partners only, eight reported exposure to both baseline and at least one new partner, while three reported exposure to new partners only. According to the classification algorithm using the clinical questionnaire, those repeat infections with no reported sexual exposure were treatment failures (n=30, 66.7%), those with any sexual exposure to baseline partners were classified as re-infection (n=12, 26.7%), and those with exposure to a new partner only were classified as new infection (n=3, 6.7%).

**Figure 1.**
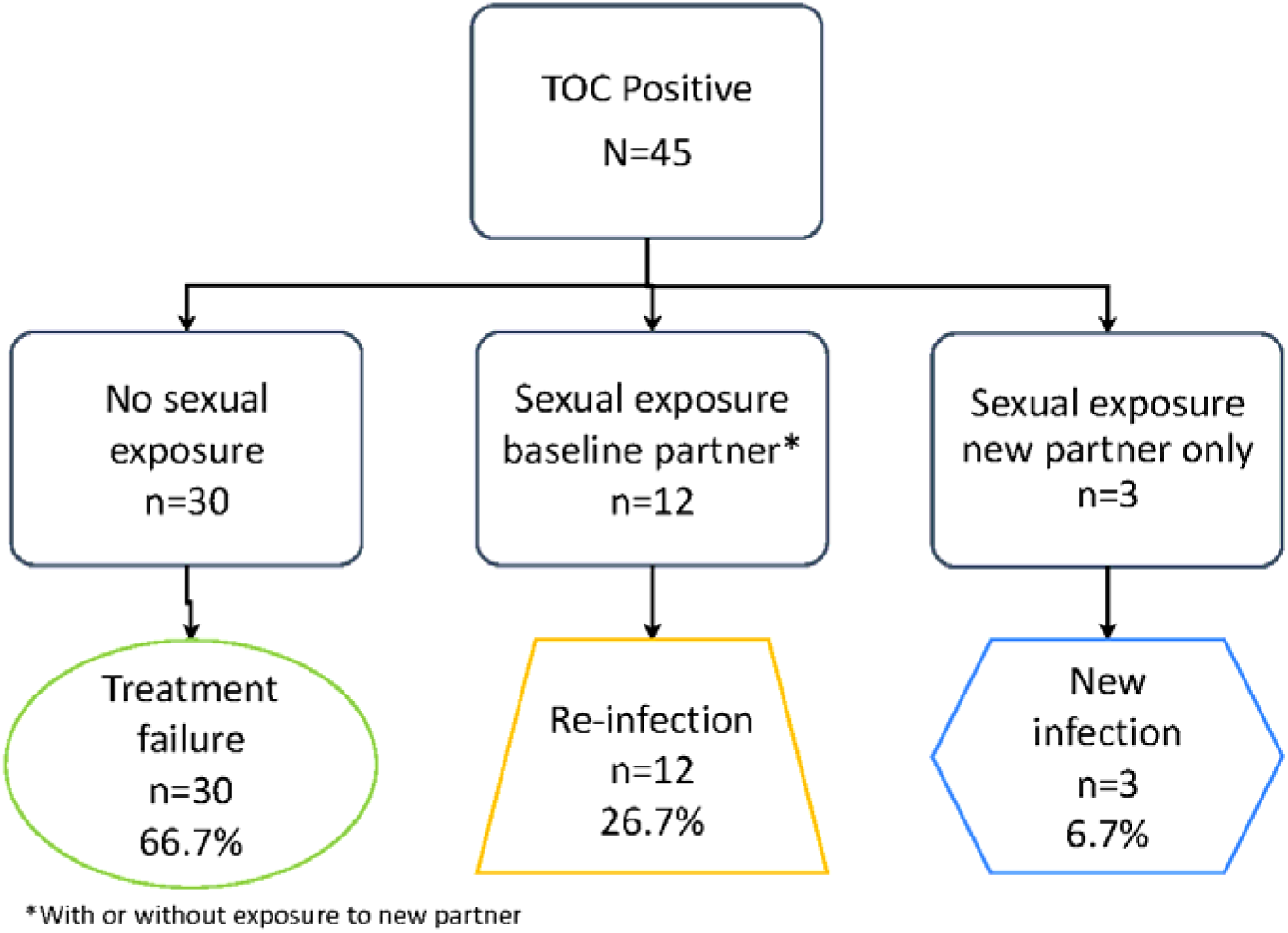
Clinical Criteria. With clinical criteria based on behavioral data alone, repeat TV infection was due to 66.7% treatment failure, 26.7% re-infection, and 6.7% new infection.

The genotype classification algorithm was defined as follows. All those with a discordant TV genotype at TOC were classified as new infection. Outcomes for concordant genotype were classified as follows: concordant genotypes and no reported sexual exposure as treatment failure, concordant genotype and exposure to a baseline partner as re-infection, and concordant genotypes and exposure to new partners only as treatment failure.

To determine the threshold of MS loci differences to define genotype discordance between baseline and TOC, we examined results at a range of cutoffs (Table 2). When ≥ 2 or ≥ 3 loci cutoffs were used, the percentages of treatment failure and new infection were highly variable. The classification was less sensitive to changes at ≥ 4 loci different or higher cutoffs, thus ≥ 4 was used to define discordance. Genotype concordance was defined as <4 MS loci different and genotype discordance was defined as ≥ 4 MS loci different.

**Table 2.** Sensitivity of *Trichomonas vaginalis* repeat infection classification to number of microsatellite loci different

|  |  | Number of microsatellite loci different |  |  |  |  |
| --- | --- | --- | --- | --- | --- | --- |
|  |  | n (%) |  |  |  |  |
|  | Clinical | ≥ 2 loci | ≥ 3 loci | ≥ 4 loci | ≥ 5 loci | ≥ 6 loci |
| Treatment failure | 30 (66.7) | 15 (30.0) | 24 (53.3) | 29 (64.4) | 30 (66.7) | 30 (66.7) |
| Re-infection | 12 (26.7) | 8 (17.8) | 8 (17.8) | 8 (17.8) | 8 (17.8) | 8 (17.8) |
| New infection | 3 (6.7) | 22 (48.9) | 13 (28.9) | 8 (17.8) | 7 (15.6) | 7 (15.6) |

Comparing genotypes from baseline and TOC, 31 of the 45 participants had genotype concordance, and 14 had genotype discordance (Figure 2). Of those with genotype concordance, 22 reported no sexual exposure, seven reported sexual exposure to baseline partner, and two reported exposure to a new partner only. Those with genotype concordance and with either no sexual exposure or exposure to new partners only were classified as treatment failure (n=29, 64.4%). Those with genotype concordance and exposure to their baseline partners were classified as re-infection (n=8, 17.8%). Among participants who had genotype discordance, four reported no sexual exposure, three reported exposure to their baseline partners, and one participant reported exposure to new partners only. All participants with genotype discordance were classified as new infections (n=8, 17.8%).

**Figure 2.**
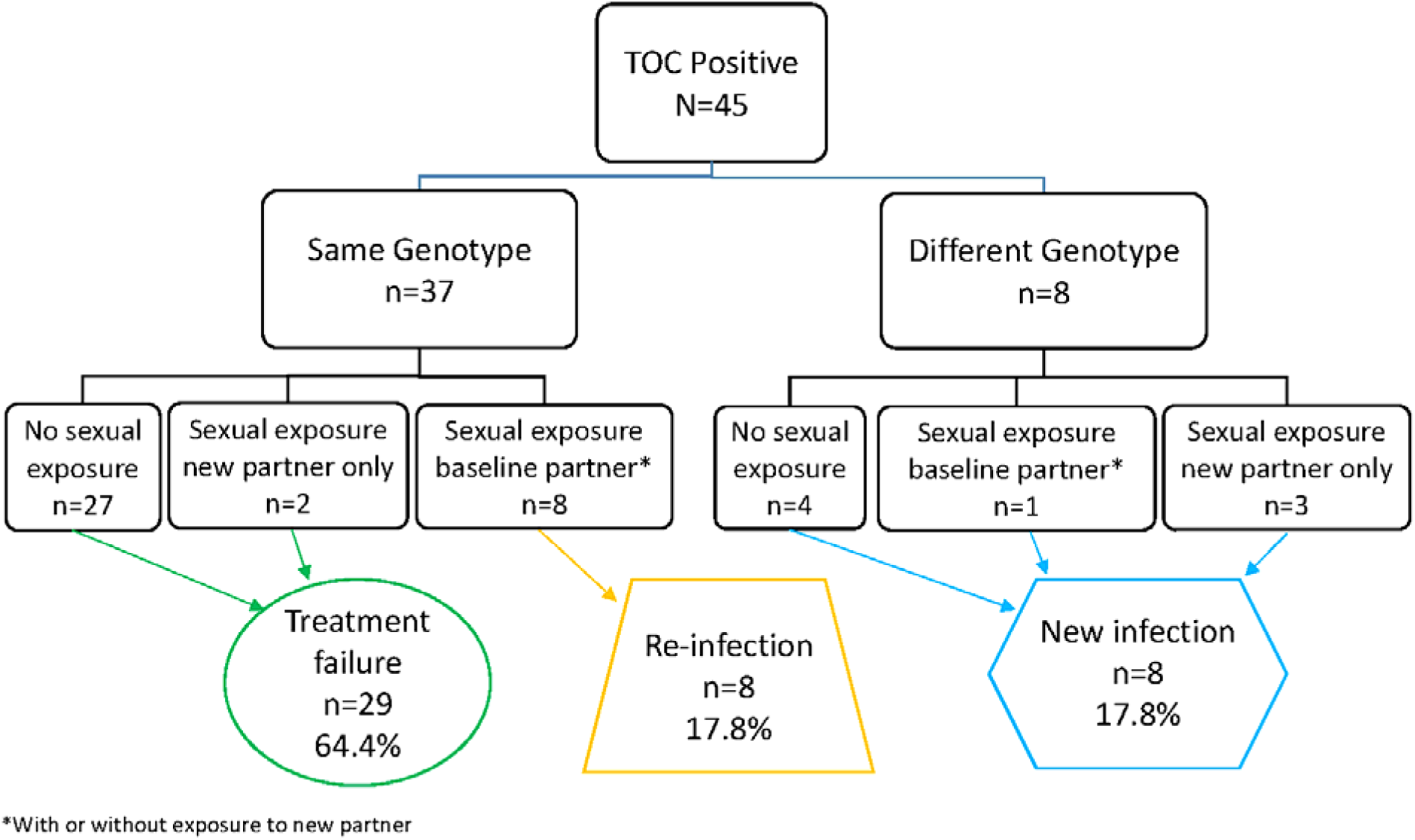
Genotype-informed Criteria. With genotype-informed criteria based on MS genotype and behavioral data, repeat TV infection was due to 64.4% treatment failure, 17.8% re-infection, and 17.8% new infection.

One re-infection and two new infections under clinical criteria were reclassified as treatment failure with genotypic criteria (Table 3). Three re-infections and four treatment failures under clinical criteria were reclassified as new infections by genotype-informed criteria. Cohen’s kappa for reliability was 0.57 (95% CI 0.36 - 0.78). The proportion of cases classified in three categories changed with the addition of genotypic criteria (Χ^2^=16.00 p=0.0011).

**Table 3.** Comparison of *Trichomonas vaginalis* repeat infection classifications

| Clinical<br>Classification | Genotype-Informed<br>n (%) |  |  |  |
| --- | --- | --- | --- | --- |
|  | Treatment failure | Re-infection | New Infection | Total |
| Treatment failure | 26 (57.9) | 0 (0.0) | 4 (8.9) | 30 (66.7) |
| Re-infection | 1 (2.2) | 8 (17.8) | 3 (6.7) | 12 (26.7) |
| New Infection | 2 (4.4) | 0 (0.0) | 1 (2.2) | 3 (6.7) |
| Total | 29 (64.4) | 8 (17.8) | 8 (17.8) | 45 (100.0) |

## Discussion

In this study, a majority of repeat infections were classified as treatment failure for both genotype and clinical criteria. With addition of genotype data to clinical assessment, there were fewer repeat infections classified as re-infections and more infections classified as new infections.

This study has several strengths. In the United States, TV prevalence is 6-10% higher in black/African American women than non-Hispanic white women [4, 5], thus participants in our study reflect the highest risk group for TV infection. They must be prioritized in interventions and policies targeted at TV, and our research provides further insight on the infection process in this population. This study was the first to evaluate the origin of repeat TV infection with genotypic criteria in women uninfected with HIV. In this population, we determined the optimal timing for NAAT testing to be four weeks post-completion of treatment [16]. Nucleic acid detected at four weeks TOC reflect ongoing infection, not residual nucleic acids from a successfully treated original infection.

While 6.7% of participants had new infections using only clinical criteria, 17.8% of participants had new infection using genotypic criteria. These new infections may come from exposure to a new infected partner, a new infection acquired by baseline partners during follow-up, or emergence of a second strain present but not detected at baseline (i.e. a misclassified treatment failure). The latter would represent an initial mixed infection initially not detected because the newly identified strain originally constituted a small minority of the organisms present. New infection may contribute more to repeat infection than currently estimated by clinical criteria. Studies which used genotype-informed classifications for repeat *Chlamydia trachomatis* infection also found an unexpectedly high percentage of new infections [11, 17]. It is possible the participants did not accurately report sexual exposure, had unknown exposure through partners, or the genotyping had limitations. Here women were interviewed using ACASI, which has been found to reduce social desirability bias [18], but reporter bias cannot be ruled out given that there is no gold standard.

Participants with any sexual exposure to a new partner are classified as new infection using clinical criteria, we used Bayesian reasoning to estimate the probability of whether the infection might be due to treatment failure given the information known about their genotype. In participants who reported only new partners, given the genotype was the same, we concluded there was a higher probability infection was due to treatment failure rather than new infection. However, it is also possible that a woman could be re-infected with the same TV strain by a new partner, especially if they all belonged to the same sexual network.

In the case of exposure to baseline partners and an organism with the same genotype as baseline, we cannot differentiate treatment failure from re-infection with MS data alone. All those with sexual contact to baseline partners are classified as re-infection, though the true reason for recurrence at TOC is treatment failure due to the regimen itself in some cases.

Understanding of the origin of a repeat TV infection can help determine the best approach to preventing these infections. Re-infection from baseline partners can be addressed through expedited partner therapy, the practice of providing medications or prescriptions to the sex partners of the index patient without requiring the partners to first visit a healthcare provider. Treatment failure and new infection, which represented 82.2% of infections in this study, can be addressed by rescreening after completion of treatment. A high percentage of treatment failure also indicates an insufficient treatment regimen. In this study, twice as many repeat infections came from the single-dose 2g MTZ regimen as the 7 day BID dose (n=30 and n=15, respectively). Two RCTs and one meta-analysis have demonstrated that multi-dose metronidazole is superior to the single 2 g dose and CDC will now recommend multi-dose MTZ treatment [9, 19-22].

In this study, the origin of repeat infection in a majority of subjects was treatment failure by either clinical (66.7%) or genotypic (64.4%) classification. The reclassification to new partner using genotypic criteria could mean that participants are underreporting exposure to new partners, original partners are infecting the women with new types, or women carry mixed infections where one genotype is cleared and another is detected. Genotyping may provide a more information with which to assess of the origin of repeat infection if used in conjunction with clinical assessment.

## Data Availability

All data produced in the present study are available upon reasonable request to the authors

## Funding

This work was supported by the National Institute of Allergy and Infectious Diseases at the National Institutes of Health [1R01AI097080–01A1].

## Conflicts of Interest

The authors declare no conflicts of interest.

## Acknowledgements

PA and WES from the CDC provided their services in kind. We thank the Data Safety and Monitoring Board, including Charlotte Gaydos, David Mushatt, Jeffery Burton, and Russell Van Dyke. We thank Caroline Deal and Hagit David from the Division of Microbiology and Infectious Diseases of the National Institute of Allergy and Infectious Diseases and DMID for their support. At University of Alabama at Birmingham we thank the clinical and laboratory staff, including Hanna Harbison, Saralyn Richter, Rhonda Whidden, Meghan Whitfield, Christen Press, Jim Alosi, Ann Dillashaw, Charles Rivers, Cheri Aycock, and Keonte Graves. At University of Mississippi Medical Center we thank Melverta Bender and Jennifer Brumfield. At Louisiana State University we thank Camille Fournet, and at the Louisiana State University Health Sciences Center we thank the laboratory staff, including Catherine Cammarata, Judy Burnett, and Denise Diodene. We also thank the data management staff at Tulane University, including Lauren Ostrenga and Scott White. We thank Grace Tooley and Jonathan Bermeo who helped with wet-lab work during their studies at NYU. The findings and conclusions in this study are those of the authors and do not necessarily represent the views of the National Institutes of Health, National Institute of Allergy and Infectious Diseases, or the CDC.

## Notes

### Competing Interest Statement

The authors have declared no competing interest.

### Clinical Trial

NCT01018095

### Clinical Protocols

https://www.ncbi.nlm.nih.gov/pmc/articles/PMC6279510/

### Author Declarations

The study received ethics approval from the institutional review boards of Tulane University, Louisiana State University Health Sciences Center (LSUHSC), University of Alabama at Birmingham, University of Mississippi MedicalCenter,JeffersonCountyDepartmentofHealth, and the Mississippi State Department of Health.

